# Automated self-service cohort selection for large-scale population sciences and observational research: The California Teachers Study Researcher Platform

**DOI:** 10.1101/2023.12.22.23300461

**Authors:** James V. Lacey, Emma S. Spielfogel, Jennifer L. Benbow, Kristen E. Savage, Kai Lin, Cheryl A.M. Anderson, Jessica Clague-DeHart, Christine N. Duffy, Maria Elena Martinez, Hannah Lui Park, Caroline A. Thompson, Sophia S. Wang, Sandeep Chandra

## Abstract

**Objective:** Cohort selection is ubiquitous and essential, but manual and ad hoc approaches are time-consuming, labor-intense, and difficult to scale. We sought to automate the task of cohort selection by building self-service tools that enable researchers to independently generate datasets for population sciences research.

**Materials and Methods:** The California Teachers Study (CTS) is a prospective observational study of 133,477 women who have been followed continuously since 1995. The CTS includes extensive survey-based and real-world data from cancer, hospitalization, and mortality linkages. We curated data from our data warehouse into a column-oriented database and developed a researcher-facing web application that guides researchers through the project lifecycle; captures researchers’ inputs; and automatically generates custom and analysis-ready data, code, dictionaries, and documentation.

**Results:** Researchers can register, access data, and propose projects on the CTS Researcher Platform via our CTS website. The Platform supports cohort and cross-sectional study designs for cancer, mortality, and any other ICD-based phenotypes or endpoints. User-friendly prompts and menus capture analytic design, inclusion/exclusion criteria, endpoint definitions, censoring rules, and covariate selection. Our platform empowers researchers everywhere to query, choose, review, and automatically and quickly receive custom data, analytic scripts, and documentation for their research projects. Research teams can review, revise, and update their choices anytime.

**Discussion:** We replaced inefficient traditional cohort-selection processes with an integrated self-service approach that simplifies and improves cohort selection for all stakeholders. Compared with manual methods, our solution is faster and more scalable, user-friendly, and collaborative. Other studies could re-configure our individual database, project-tracking, website, and data-delivery components for their own specific needs, or they could utilize other widely available solutions (e.g., alternative database or project-tracking tools) to enable similarly automated cohort-selection in their own settings. Our comprehensive and flexible framework could be adopted to improve cohort selection in other population sciences and observational research settings.

## Introduction

Observational research using real-world data makes vital contributions to biomedical research (2). Large cohort studies of volunteers whose data are tracked and aggregated for research play an especially important role and often become community resources (26). The largest cohorts, including the NIH All of Us Research Program (2), UK Biobank (12), and Million Veteran Program (21), can include hundreds of thousands of participant partners (i.e., volunteers), last for decades, and support a wide range of future research projects (34) and broad data sharing (32).

Individual research projects rarely require all the data a large cohort has assembled. “Cohort selection” refers to the process of generating project-specific datasets that give researchers what they need while protecting participants’ privacy and confidentiality. Cohort selection includes specifying the study design; applying eligibility, inclusion, and exclusion criteria; operationally defining key study parameters and endpoints; and choosing specific covariates. Growing use of Electronic Health Records (EHRs) (30) and research data warehouses and repositories (22) relies on cohort selection. The CONSORT (4) and STROBE (35) statements recommend that reports of study results thoroughly document the cohort selection process.

When data are private or proprietary, data providers can spend considerable time and energy helping data requestors understand the data, optimize requests, and perform cohort selection (26). Even with modern computing infrastructure, manual cohort-selection takes too long and cannot scale (17); cohort selection is often a bottlenecking event. Even after deploying a cloud-based data commons specifically designed to improve data access and sharing (19), our prospective California Teachers Study (CTS) cohort (5) struggled with manual cohort selection.

Other cohorts (37) and enterprises (17) provide self-service query tools. We sought to enable complete and comprehensive self-service cohort selection, including data delivery. This report describes our development and deployment of the CTS Researcher Platform, an innovative tool that empowers researchers everywhere to query, choose, review, and automatically and quickly receive custom data, analytic scripts, and documentation for their research projects.

## Materials and methods

### The California Teachers Study (CTS)

The CTS is an NCI-funded, multi-site prospective cancer epidemiology cohort (CEC) study (5). It began in 1995-1996, when 133,477 adult women completed a survey and consented to future data collection and use of their data for research (9). Participants completed up to five follow-up surveys that covered diverse health, lifestyle, and environmental exposures (10). Ongoing annual data linkages with the California Cancer Registry (CCR), Department of Health Care Access and Information (HCAI; formerly Office of Statewide Health Planning and Development (OSHPD)), and Department of Public Health Vital Records (CDPH-VR) have identified over 36,000 participants with cancer, over 108,000 participants who were hospitalized, and over 38,000 participants who died during follow-up (6). Linkages with the Centers for Medicare and Medicaid Services (CMS) identified detailed provider and claims data for over 98,000 participants. With additional biospecimens, biomarkers, and linked geospatial data, the CTS’s survey and real-world data can enable hundreds of potential research projects (8).

### Ethics

Based on the study invitation they received, participants who completed the baseline CTS survey were considered to have provided informed consent, with a waiver of written informed consent based on return of the completed baseline survey. CTS data are under controlled access to protect participants’ privacy and confidentiality. The first and last completed baseline surveys were received on Oct. 27, 1995, and Aug. 20, 1999, respectively.

### CTS data environment and previous cohort selection methods

Until 2015, we used entirely manual cohort selection: data managers received individual requests, worked with requestors, and created datasets with accompanying dictionaries, using locally stored files and desktop software. In 2016, our data commons (19) replaced those silos and brought users, data, software, and tools together in one secure shared environment that includes 1) a data warehouse with data marts designed for analysis; 2) file and extract-transform-load (ETL) servers; 3) software, tools, metadata, and documentation; and 4) a remote desktop environment that serves as the collaborative workspace.

### Cohort selection: Still a rate-limiting step

The CTS data commons (19) did not eliminate manual cohort selection. We standardized the process by asking users to specify their inclusion/exclusion criteria, endpoints, covariates, and other details using drop-down menus and open-ended text in Excel worksheets. CTS data analysts then manually inserted those choices into SAS Proc SQL templates (over 90% of projects and analysts used SAS (Cary, NC) software) to join data from the CTS warehouse and/or marts. Researchers then used those customized templates to call and analyze their project-specific data in our secure CTS workspace. Even with cloud-based data, data calls, and workspaces, manual entries by researchers and manual customization of data calls by the CTS team were unsustainable and unscalable.

### New challenge: Self-service cohort selection at scale

We needed to eliminate manual cohort selection. Instead of giving their choices to CTS analysts, researchers should be free to directly interact with CTS data, automatically apply their choices, and generate their own data. Our data commons already provided a secure workspace with the data, tools, and documentation users needed (19), but it needed three new features: 1) <u>flexible, comprehensive, and robust menu-driven workflows</u> that include the full range of cohort selection choices without limiting researchers’ options; 2) a <u>web-based application</u> for applying those choices to our data source; and 3) <u>workflows for automatically generating the required</u> <u>deliverables</u>, including datasets, data dictionaries, and analysis scripts.

### Workflows: Robust yet flexible

The CTS is primarily a “risk” CEC, rather than a “survivorship” CEC (23); this shapes our cohort selection choices. Using our existing CTS templates, we identified the full range of potential study designs (case-control, cohort/time-to-event, cross-sectional), analytic outcomes (incident cancer, mortality, hospital- or ICD-based outcomes/phenotypes), and exposure data (self-report from surveys, geospatial data, biospecimen-based data) that are typical of risk cohorts. Our initial solution focused on the most common type of CTS project to date: a cohort design with an individual cancer type as the analytic outcome and self-reported survey data as the main exposures (8).

We articulated detailed user stories (38) to capture design requirements across the entire process. These included the ability to select multiple outcomes and multiple exposures; choose covariates individually or by hierarchical categories; set specific start-of-follow-up and end-of-follow-up dates; establish inclusion and exclusion criteria; apply analytic censoring rules; and review real-time dashboards that displayed frequencies based on users’ inputs. Another essential user requirement was the ability to revise any individual component of the cohort selection—e.g., to add independent variables or modify censoring criteria—during a project without requiring users to completely start over. We identified the types of deliverables users would need (e.g., custom data dictionaries with only the covariates they had selected; a summary of their design decisions, etc.). We decided to configure datasets that could be analyzed using open-source (e.g., Posit, formerly R Studio) or commercial (e.g., SAS) analysis software packages. We managed user stories and project management via smartsheet.com, mock-ups via figma.com, and communication and collaboration via Slack.

We also identified combinations of design choices that initially seemed too complex to automate, such as idiosyncratic matching criteria for controls in nested case-control projects and clinical phenotypes based on complex combinations of ICD codes and hospitalization patterns. We excluded those workflows from the initial solution.

### Source data: High-performance data management from multiple domains

CTS data linkages update cancer, hospitalization, and mortality data annually (19), but survey data are static. This allowed us to extract data from our warehouse and avoid having the self-service tool directly query our data marts. We also considered missing-by-design data: in large prospective studies, study censoring (e.g., participants who die do not complete subsequent surveys) and rare outcomes (e.g., low frequencies of multiple-primary cancers) create valuable information but are inefficient from a traditional SQL database perspective, because they create large numbers of columns, but many of those columns contain empty cells. To increase flexibility, scalability, and speed, we added a columnar (column-oriented) online analytical processing (OLAP) database management system designed for high-performance (ClickHouse, Redwood City, CA) as the data source that would be available to the self-service web application.

We wrote a script that extracts data from our data warehouse from three domains—participants, cancers, and surveys—into a large and wide OLAP database. Participant data included essential characteristics (e.g., date of birth, date of death, cause of death, race/ethnicity, vital status) and follow-up information (e.g., dates of study enrollment, follow-up surveys, last follow-up, etc.). Cancer data included detailed site, stage, grade, diagnosis date, etc. for all cancers during follow-up. Survey data included approximately 1200 covariates (i.e., columns) from the CTS questionnaires, with at least one column for every question, plus other existing derived covariates derived (e.g., body mass index (BMI) based on self-reported height and weight). All questionnaire covariates were previously tagged to facilitate identification by question number, questionnaire section, or questionnaire number (6). The workflow for creating and updating this presentation database leveraged the efficient schema of our data warehouse to generate individual participant records across all domains.

### Data and code availability

All CTS data and analytic code are available in the CTS Researcher Platform (7), which is also available via the “For Researchers” tab on the CTS website (5). CTS data are not publicly available because they include extensive identifiable, sensitive, and confidential information, but any researcher who agrees to protect and use CTS data responsibly for research can access all CTS resources through the Researcher Platform (7). The underlying code for the self-service cohort-selection application is available upon request via the Researcher Platform (7).

## Results

### Developing the web application: User-friendly menus and interactive visualizations

User stories and the potential study design and analysis choices drove the web application development. We wanted to emulate the user-friendly menus and buttons in tools such as the NCI SEER*Explorer Application (SEER*Explorer) (27). In our experience, all researchers who ask to use data from cohorts like the CTS for their projects understand the basics of cohort selection. We designed the application for novice users who did not have any prior experience analyzing CTS data; i.e., the application needed to walk users through every step of the process in easily understandable ways.

We identified six steps in the cohort selection process and created a sequential task-based page in the application for each step: 1) select cancer endpoint; 2) select start of follow-up; 3) select censoring rules; 4) select questionnaire data; 5) enter biospecimen data; and 6) review summary. The application includes drop-down menus and search functions to help users choose their cohorts and data. Users can choose cancer endpoints by ICD-O-3 site or SEER Site Group Recode values (31). Additional choices can be made by histology codes. Users can start their follow-up at any of the dates that participants completed a CTS survey or enter a different start-date in the form of MM/DD/YYYY. Users can choose whether to censor participants who develop cancers other than their analytic endpoint. When users choose breast, uterine, or ovarian cancer endpoints, censoring rules automatically incorporate censoring at the date of bilateral mastectomy, hysterectomy, or bilateral oophorectomy, respectively; all are available through the CTS’s linked hospitalization data. As users choose their data, interactive visualizations display and update, in real-time, frequencies and distributions of their choices.

The application automatically saves users’ interim progress and allows all members of a project team to make or modify choices. Each page includes a “back” button that returns users to the previous page, where choices can be modified. Figure 1 shows screenshots from a sample cohort design with a cancer endpoint and survey-based exposures.

**Fig 1.**
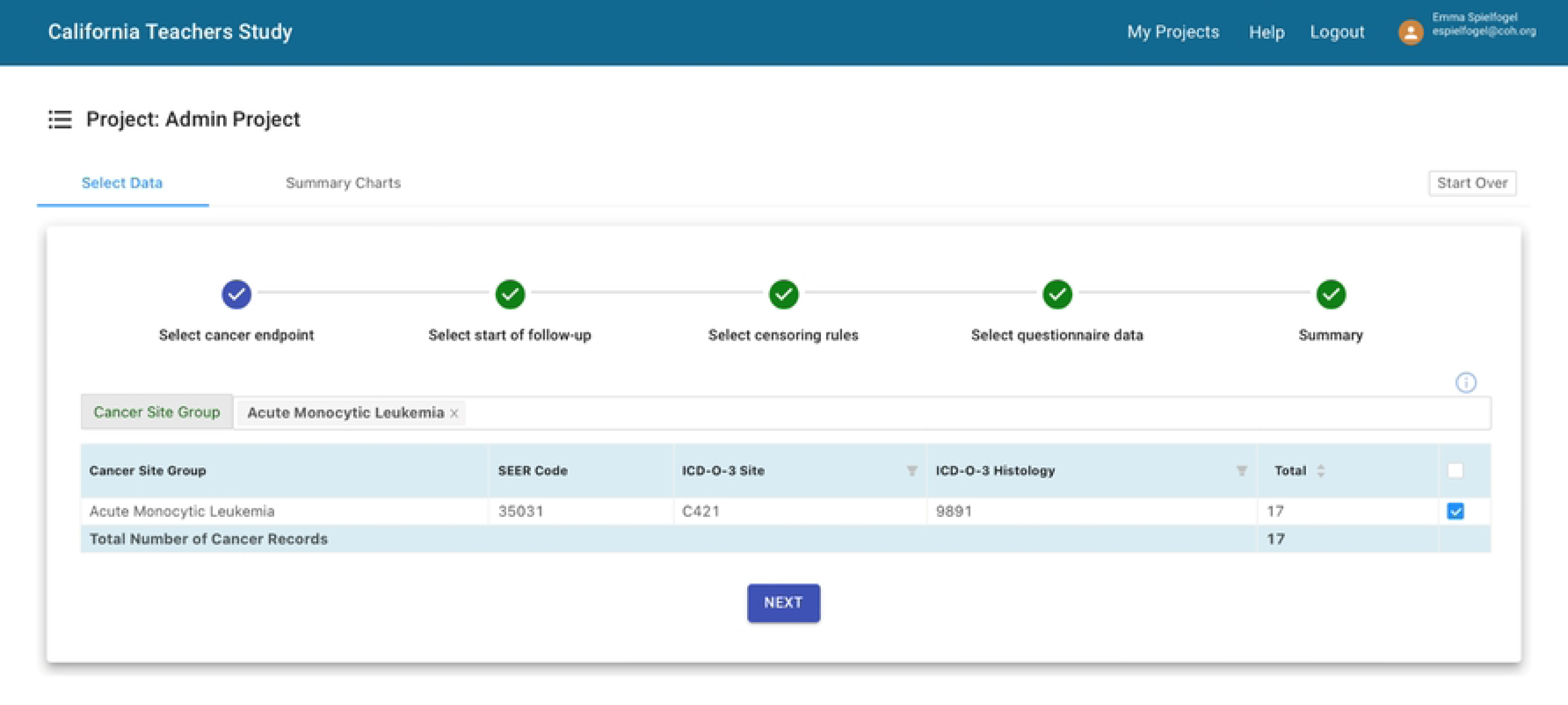
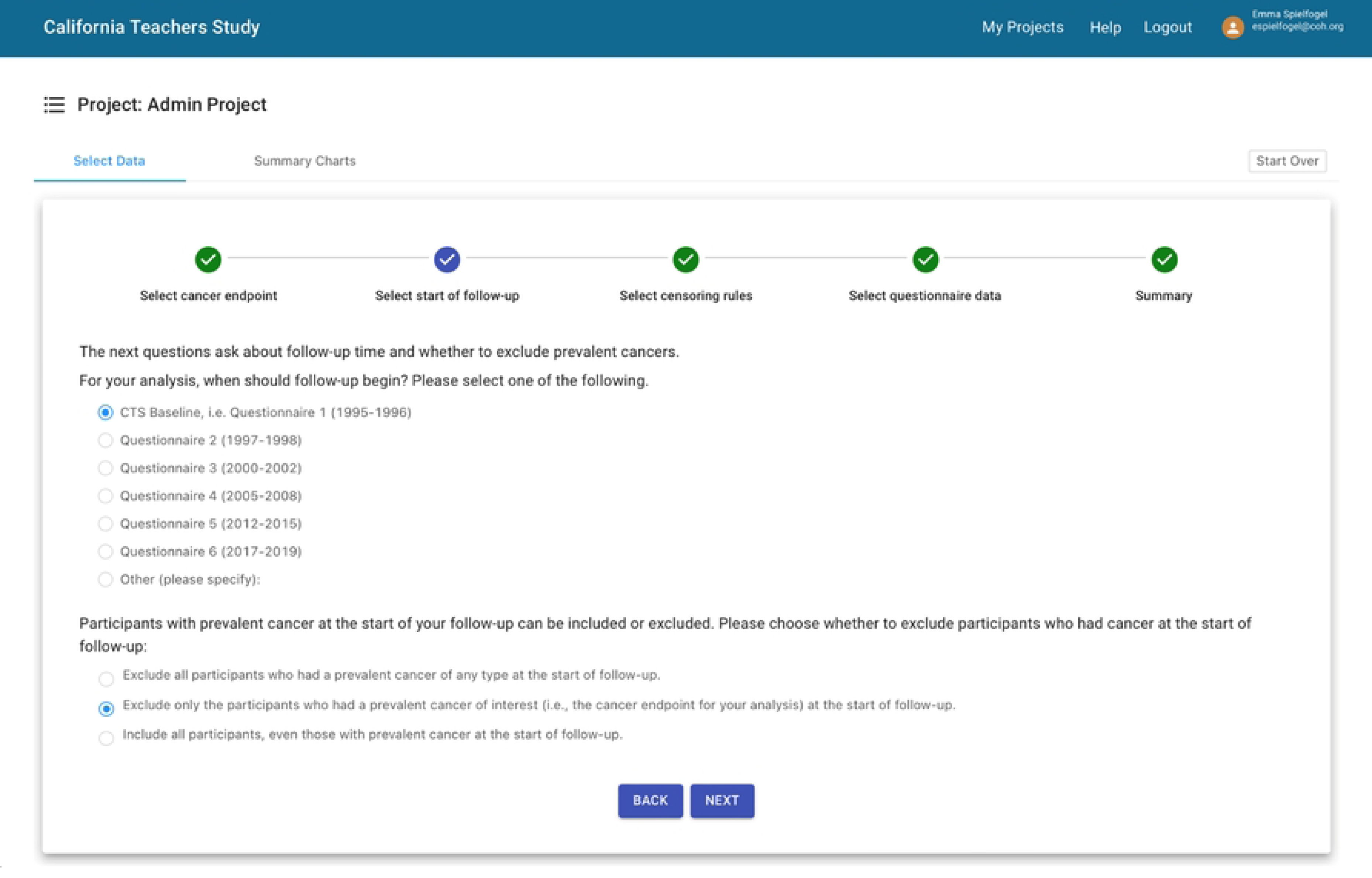
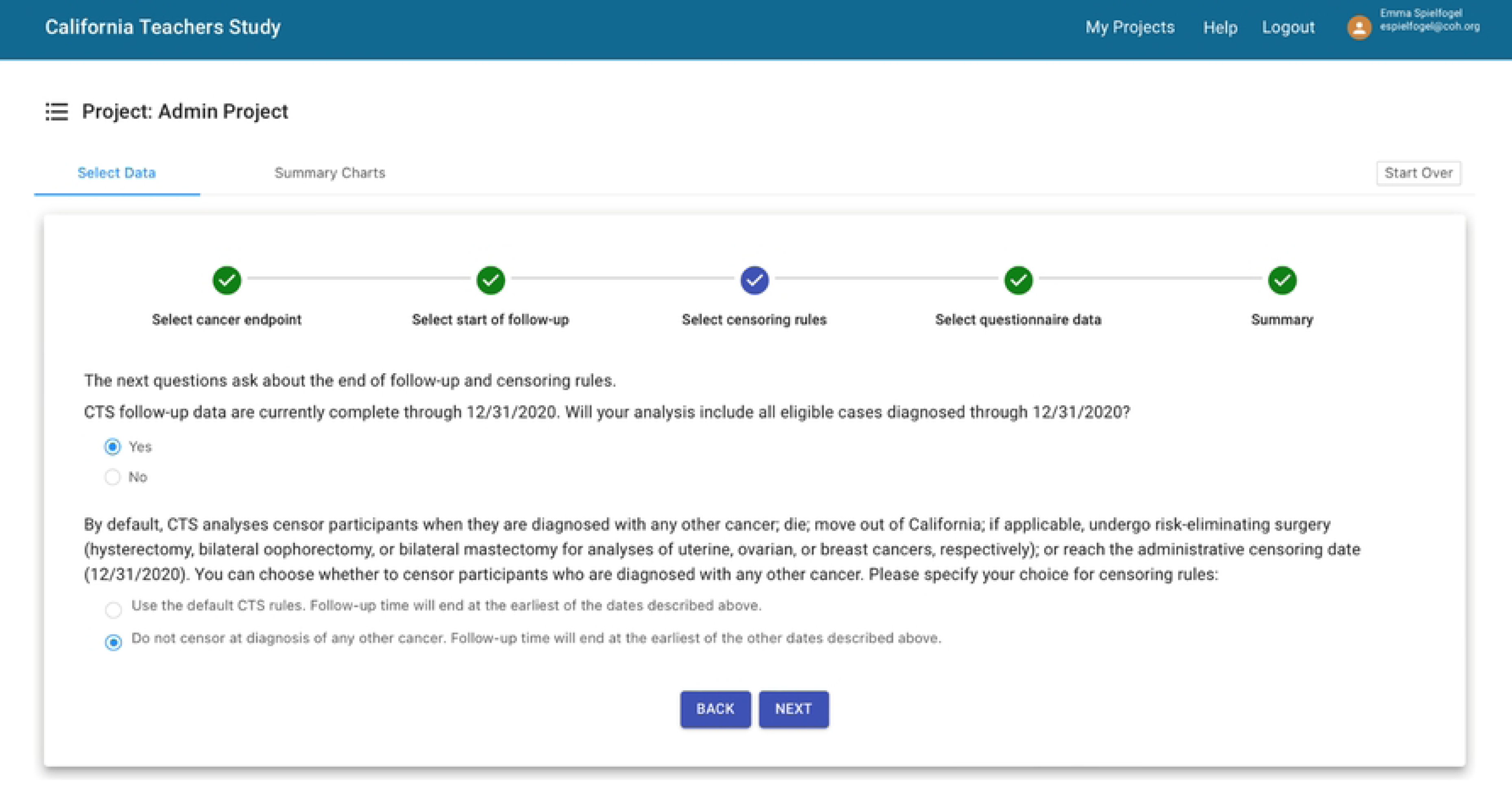
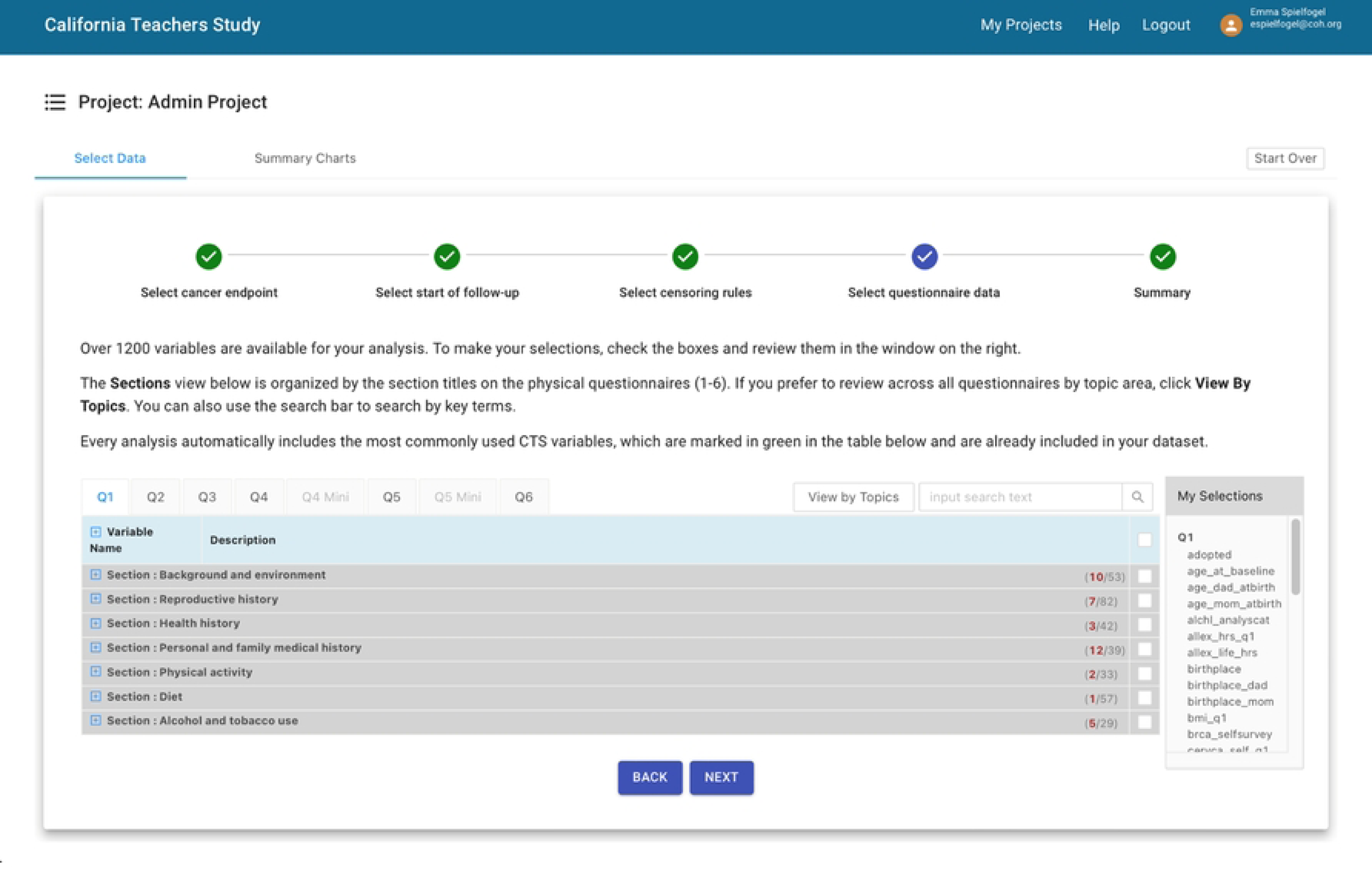
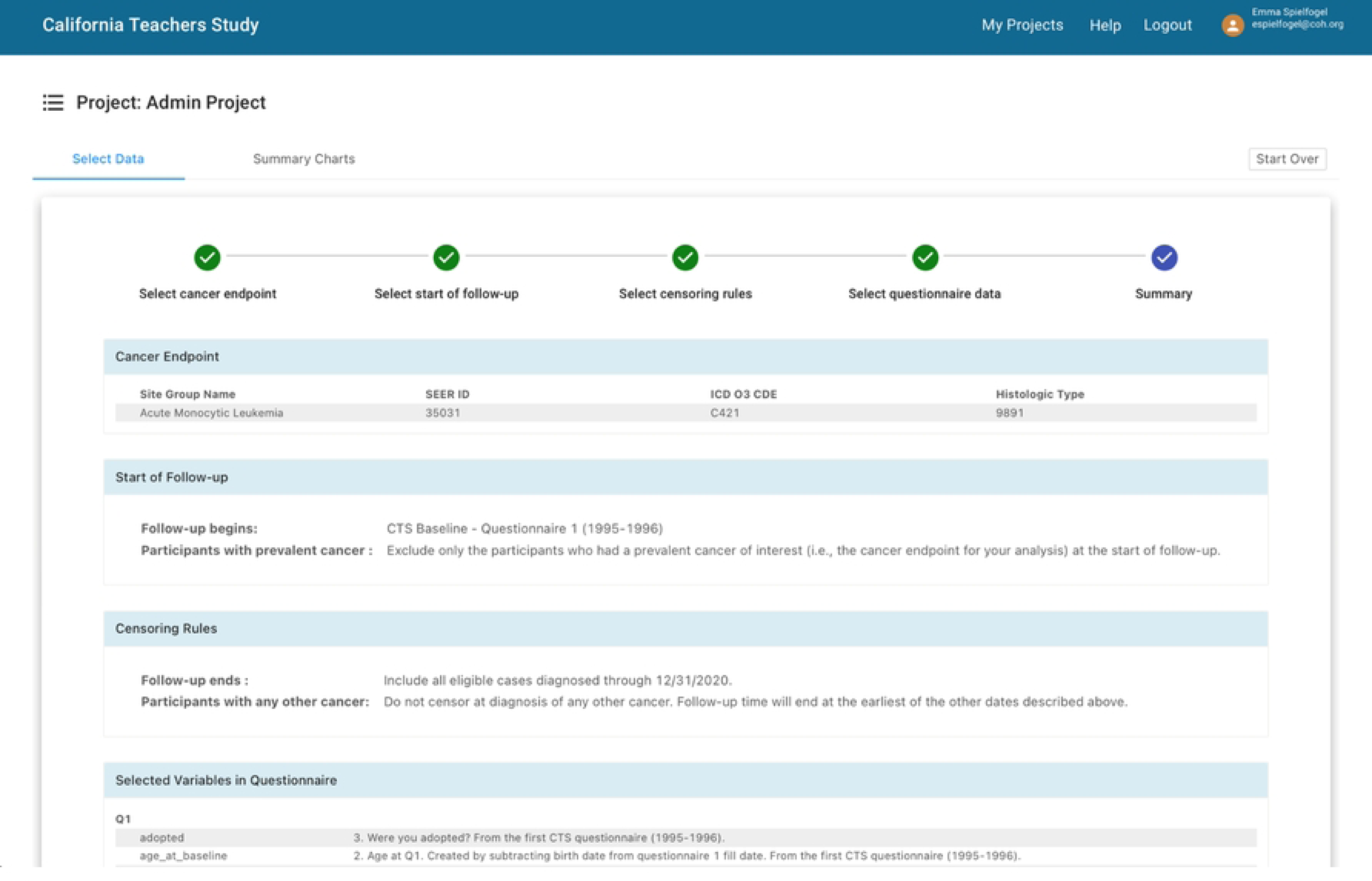
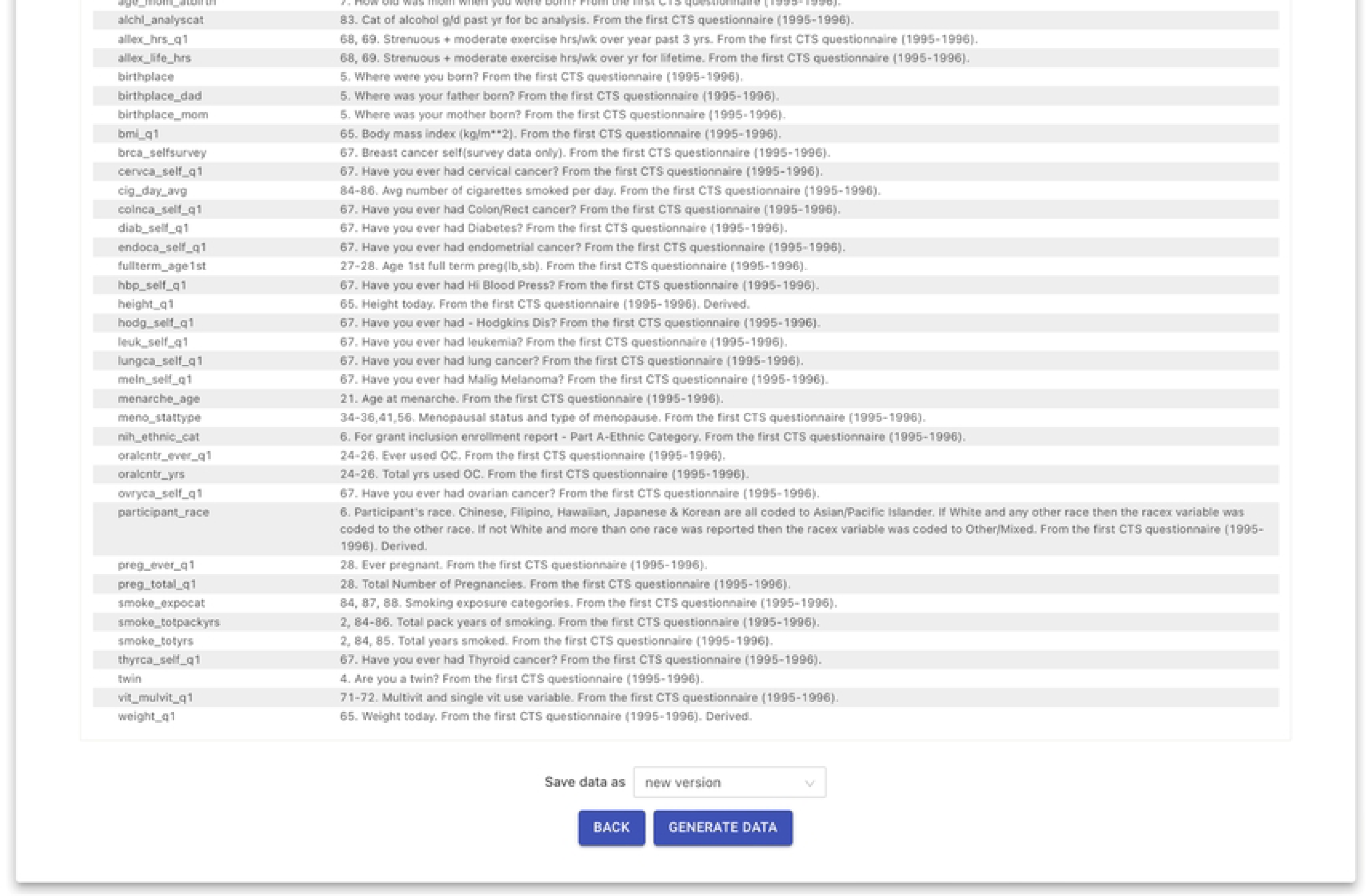
Screenshots of the self-service cohort selection application for a cohort analysis with a cancer endpoint and survey-based exposure data. (a) Select cancer endpoint. (b) Select start of follow-up. (c) Select censoring rules. (d) Select questionnaire data. (e) Summary.

### Deliverables: Immediate access to data and continuous CTS support

The final screen includes a “Generate Data” button. When users click that button, the application saves all of the inputs and generates six deliverables: 1) a custom *.csv dataset based on the user’s choices; 2) a SAS-specific formats file matching the custom *.csv dataset (SAS is the primary software used in the CEC community); 3) a SAS data call that brings the custom *.csv dataset into SAS, using the formats file to automatically apply the appropriate data formats for analysis; 4) an Posit (R Studio) script that reads the custom *.csv dataset into a new R session for analysis; 5) a custom data dictionary that includes all of the covariates selected and omits all CTS covariates that were not selected; and 6) a PDF summary of all the cohort selection choices. The dataset and formats files are automatically written to a read-only directory and the data calls, dictionary, and summary file are automatically written to that project’s dedicated directory, all within the CTS’s remote desktop environment (19).

Generating these deliverables typically requires less than 30 seconds; users essentially get immediate access to the data, tools, and documentation they need to analyze their data. Writing datasets to a read-only drive facilitates data governance, data lineage, and version control, and it preserves data fidelity for every output dataset back to the CTS data warehouse. Writing the data calls to project-specific workspaces gives researchers complete control and flexibility over what they do with their code, scripts, and analytic methods. Because all deliverables reside in the CTS’s shared workspace (19), our CTS team can assist, troubleshoot, or collaborate in real-time with any researcher on any part of any project.

### Other essential components: User accounts, project tracking, and integration

When researchers sign up for a user account on the CTS website (7), their account details are tracked in the CTS’s Salesforce organization. Salesforce also serves as the back-end of the “For Researchers” page on the CTS website, where researchers can propose, submit, and track their projects. Both our CTS team and researchers can see status updates as projects move through the research lifecycle. This tracking also enables researchers to automatically receive access to the cohort selection web application as soon as their project meets required IRB and approval criteria (7). We use smartsheet.com to track additional details of every project (Fig. 2).

**Fig 2.**
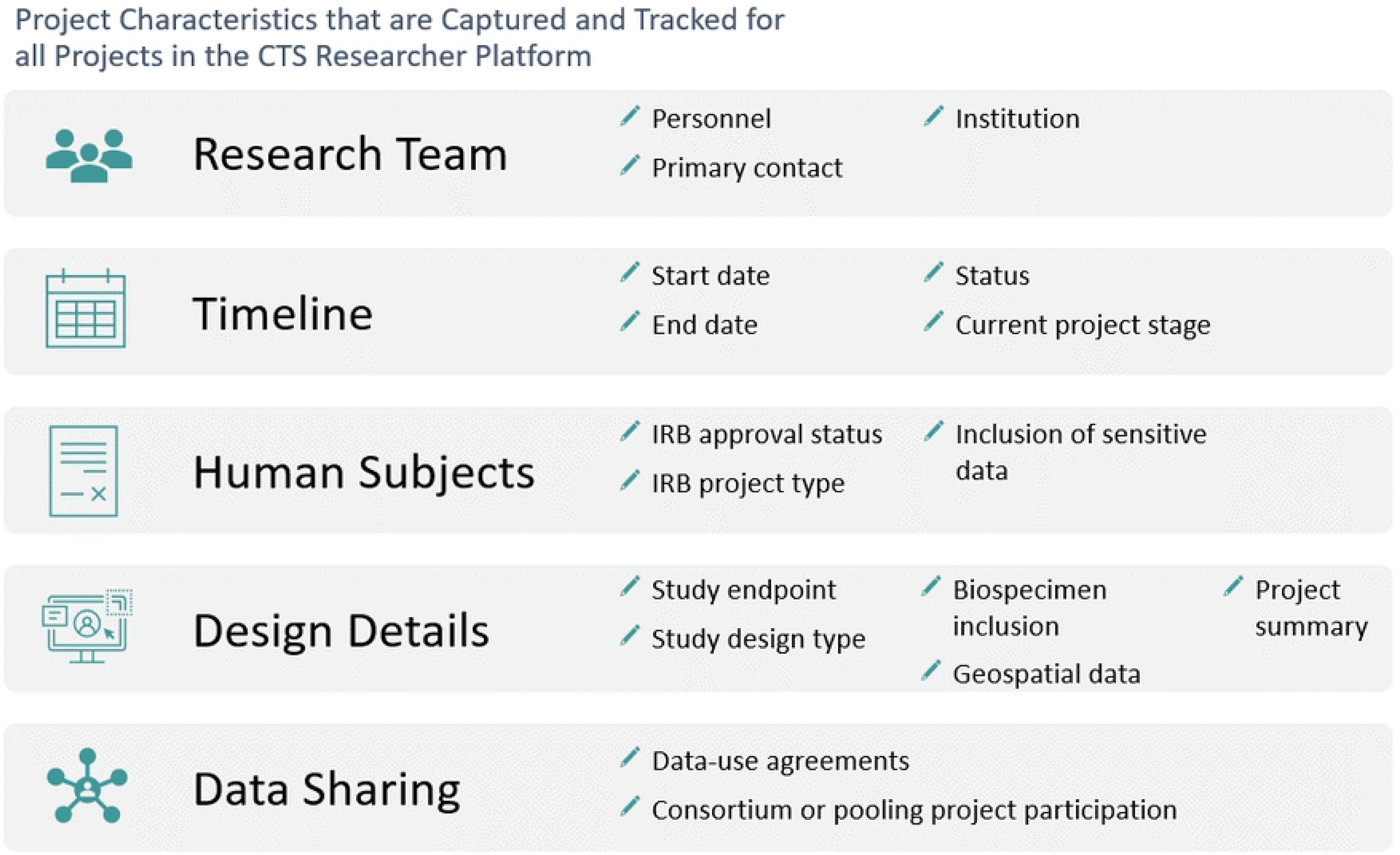
Project characteristics that are captured and tracked for all projects in the CTS Researcher Platform.

Researchers access the web application through the CTS website (5), but the web application and column-oriented database are hosted within the San Diego Supercomputer Center’s (SDSC’s) secure Sherlock environment (29). The web application bidirectionally integrates the active directory (AD) with our secure remote desktop environment, where all user accounts have role-based permissions (19), with our Salesforce organization, where projects and project status are linked to individual users. Figure 3 provides an overview of the Platform.

**Fig. 3.**
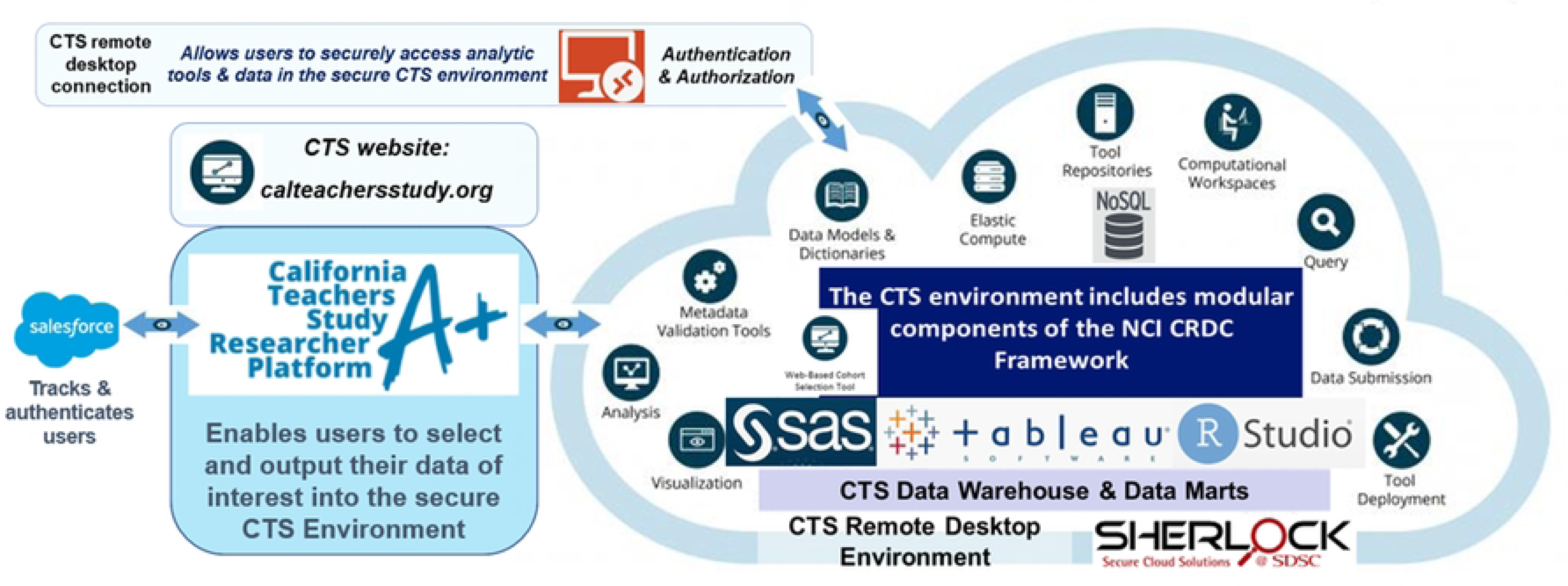
Overview of CTS Researcher Platform and integration with secure CTS environment hosted at SDSC Sherlock.

### Data scope: Standardized and customized

All datasets automatically include 62 essential covariates (e.g., dates of birth, death, and baseline survey; BMI; smoking status; etc.). Instead of requiring users to make every decision from scratch, the application provides default choices on key analytic decisions, (e.g., exclude participants with prevalent cancer), while also allowing researchers to make alternative choices.

Some complex data (e.g., geospatial-based exposures, food frequency questionnaires, etc.) were excluded from the initial database that the application uses. When a project requires those data, or if a user wants to bring their own data into a project, our CTS Research Data Steward (E.S.) uses data-call templates to deposit the needed data excerpts in the project team’s read-only directory. The excerpts use the same universal data key to facilitate easy and immediate joins for any additional or custom data; users also receive updated standard CTS code to join their custom data.

### Platform: Initial launch in 2021 and ongoing improvements

Design, development, testing, and integration took six months. After a soft launch and additional refinements (24), we launched the full platform in March 2021. Table 1 shows how cohort selection has evolved since the CTS began.

**Table 1.** Evolution of cohort selection methods and procedures in the CTS from its beginning, in 1995-1996, through the CTS Researcher Platform.

|  | Original CTS Data Strategy<br>(1995 – 2016) | Initial CTS Data Commons<br>(2016 – 2021) | CTS Researcher Platform<br>(2021 – present) |
| --- | --- | --- | --- |
| **What are the data sources?** | Locally stored SAS datasets | CTS data warehouse (DW) & data marts | NoSQL database extract from DW |
| **Who makes choices?** | CTS data managers at CTS sites | CTS-wide data analyst | Researchers who request data |
| **Who writes/edits code?** | CTS data managers at CTS sites | CTS-wide data analyst | N/A; web application replaced code |
| **Who executes code?** | CTS data managers at CTS sites | CTS-wide data analyst | Researcher clicks "Generate Data" button |
| **Where is code stored?** | Locally at CTS sites | Secure shared CTS-wide workspace | N/A; web application saves researchers' choices |
| **How are data generated?** | Local CTS manager manually creates dataset | CTS analyst runs template | Web application automatically creates data |
| **Where are data delivered?** | Local CTS manager sends data to researcher | CTS analyst deposits in project-specific folder | Web application automatically deposits data |
| **How are projects tracked?** | Manually at individual CTS sites | Manually within shared workspace | Integrated CTS website & Salesforce |
| **How are changes managed?** | Manually by local CTS data managers | Manually within shared workspace | Automated version control |
| **How much time is required?** | Weeks to months | Days to weeks | As little as 5 minutes |

The application initially supported analytic projects with incident cancer as the primary endpoint. It now also supports cohort selection for ICD-based mortality and hospitalization-based phenotypes (6). For these endpoints, a simpler query lets users select the covariates they need after skipping the cancer-related questions. Users enter their endpoint information (e.g., specific ICD codes or other requirements, such as length-of-stay) in text-based forms and then click “generate data” to create the deliverables described above. This simpler query achieves two goals. One, it often produces complete data for mortality projects, because date and cause of death are automatically included in all datasets. Two, it lets users immediately begin analyzing covariate data for hospitalization projects that require additional work by the CTS team and/or researchers to generate clinical endpoints. ICD-based phenotypes and inclusion/exclusion criteria from real-world hospitalization claims data can be complex; the application standardizes those decisions by asking users to describe their operational definition for each phenotype; multiple concurrent phenotypes are allowed. Endpoint data are then deposited, with accompanying code to join those with the output of the query, as described above.

As of Dec. 2023, 32 projects with 56 total investigators have used this application (Figure 4). Ten projects were led by students or trainees in academic programs; all generated results and internal presentations. Five projects are part of multi-study consortia. Five projects produced published or submitted manuscripts; almost all the others are still analyzing data.

**Fig. 4.**
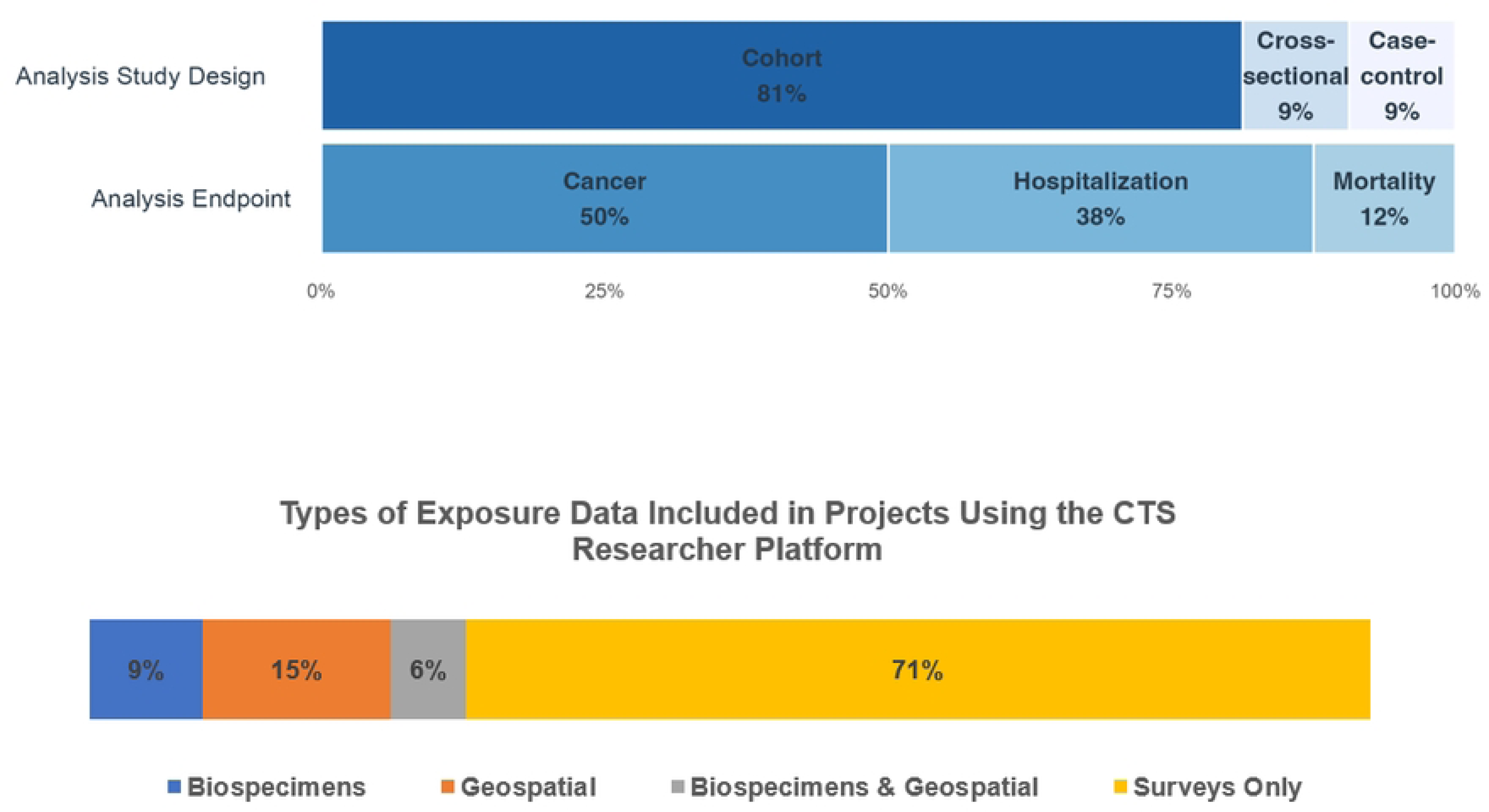
Distribution of study designs and analytic endpoints in CTS Researcher Platform projects to date.

Most projects have chosen a cohort design with cancer endpoints and survey-based exposure data, and these projects required no help from our CTS team. Projects with hospitalization-based endpoints typically require some input from our team because of the complexity of the hospitalization data. However, all those projects received complete, timely, and analysis-ready phenotype data as part of their deliverables, even when projects included multiple phenotypes. In our experience, all users have been able to independently navigate the start and stop dates, censoring decisions, and covariate selection section of the cohort-selection application.

## Discussion

Cohort selection presents significant challenges for clinical trials, cohort studies, disease registries, disease networks, enterprise-wide clinical data, and data repositories. For data providers and data requestors (26), identifying the right patients or research volunteers and then selecting the right data for those cohorts often bottlenecks the Research Data Management Lifecycle (25). Cohort selection also encounters a negative feedback loop: larger and more diverse data resources can support a wider range of research projects, but the difficulty of cohort-selection increases as the breadth, depth, and complexity of the data sources increases.

Cohort selection today usually occurs one of two ways. In one, researchers submit requests to providers, who then manually curate and deliver output. Most NCI-funded cancer epidemiology cohort (CEC) studies do this; our CTS used this approach for twenty years. This method often relies on labor-intense manual workflows, requires significant back-and-forth between investigators, and cannot scale to meet contemporary data-sharing requirements. In the other approach, providers make large source datasets available for exploration, query, and selection. This appears to be more common for enterprise-wide data providers, including electronic health records (EHRs), but also typically requires manual and project-specific assistance (13). Even forward-looking and innovative enterprise-wide query approaches, such as the Duke University Enterprise Data Unified Content Explorer (DEDUCE), struggled to provide service and data at scale (17). Two recent reviews described the challenges associated with the preliminary step of leveraging data to identify “computable clinical” (15) or “digital” (11) phenotypes and concluded that new, more efficient, and automated approaches are needed to accelerate research.

We developed a novel self-service cohort selection approach designed to eliminate manually curated datasets. We configured widely available products and software—from Microsoft Windows, Salesforce, smartsheet, and ClickHouse—and developed one new custom web application. Our integrated platform empowers users to choose and automatically receive the data and documentation they need to conduct their research; facilitates efficient collaboration and sharing; and enables us to fully track, manage, and support every user, team, and project.

Because no two cohorts are ever identical, cohort selection is typically cohort-specific. Nonetheless, our automated approach has potential for broad reusability. Regardless of where or how cohort selection occurs, it entails the same fundamental components. Cohort selection in time-to-event analyses must operationally define clinical endpoints, specify follow-up intervals, determine censoring rules, establish inclusion and exclusion criteria, and choose exposure and covariate data. Cohort selection for cross-sectional analyses requires three identical steps: define clinical endpoints, establish inclusion and exclusion criteria, and choose exposure and covariate data. We designed our approach around these common steps that are reusable across different cohort selection settings. This modular approach to cohort-selection workflows enabled us to efficiently expand our platform’s scope from just cancer endpoints to also hospitalization and mortality endpoints, while reusing other components.

The long track record of CECs successfully sharing their data in consortia, such as the NCI Cohort Consortium (32), denotes the broad potential applicability and reusability of our cohort selection approach. Dozens of CECs worldwide (20) regularly harmonize and share individual-level data for consortia projects. In those projects, every participating CEC performs cohort-selection on its data using a set of common criteria established by the consortia. The source data in each cohort are similar enough that they can be harmonized for individual-level pooled analyses (i.e., rather than meta-analyses). When similar yet independent cohorts can all execute a common cohort selection workflow to yield interoperable data that are harmonized and pooled at the level of individual participants, then the upstream cohort selection process is inherently standardizable.

Data providers’ environments, architectures, and strategies affect cohort selection. Recent papers (1,13,16–18,36) describe different approaches for leveraging research data warehouses, data marts, and repositories for research. Before developing our Researcher Platform, we used a combination of spreadsheets and SQL templates to select cohorts directly from our CTS data warehouse and data marts. We added an OLAP database as the data source for our cohort-selection application both to accelerate performance and to simplify the development of the application. Designing the application to query data from this middle layer, rather than directly from our data warehouse and data marts, also simplified our data governance strategy, because it created a buffer between CTS users and CTS source data (19). This modular approach to our solution architecture—i.e., an online database, a project tracking platform, a web-based project management tool, and a custom-built web application—can also be replicated because numerous existing tools can perform these tasks.

Two nationwide cohorts, the NIH All of Us Research Program and the UK Biobank, recently launched research platforms that make more of their data and resources FAIR (Findable, Accessible, Interoperable, Reusable). The UK Biobank initially avoided the cohort selection problem by allowing users to download source data. As the UK Biobank grew, that unsustainable strategy, which also required over a year to deliver data (14), gave way to a centralized research platform that includes analytic capabilities. The UK Biobank Research Analysis Platform (RAP) (33) utilizes a DNANexus platform that offers a variety of tools, software, and options (e.g., Spark SQL, JupyterLab, Jupyter notebooks) that researchers can use to configure and analyze data. The NIH’s All of Us Research Hub (3) includes a Data Browser for publicly available data and a Researcher Workbench for controlled-access “Registered Tier” data. This Workbench splits cohort selection into two steps: the “Cohort Builder” uses inclusion and exclusion criteria to identify a population, and the “Dataset Builder” chooses data for that cohort. Users with R or Python experience can then analyze those data in Jupyter Notebooks (3). As nationally supported cohorts, both the UK Biobank and All of Us operate at scales much larger than an individual cohort like the CTS. Within our CEC community, Python and SQL skills are not yet commonplace on research teams, and this influenced our decision to create a new point-and-click web application as the primary user interface for CTS cohort selection. Despite those differences, the NIH All of Us Researcher Workbench, the UK Biobank RAP, and our CTS Researcher Platform provide three distinct examples, at different scales, of successful web-based cohort selection.

These three first-generation cohort-selection tools reveal common themes. Robust processes and comprehensive workflows, even for historically open-ended tasks like cohort-selection, pave a path from manual methods to scalable self-service. For cohort selection, dividing the overall process into more modular units, whether the Cohort and Data Builders of the Researcher Workbench or the approach our CTS Research Platform took, works. These concepts align with emerging best practices for self-service tools more broadly: build data culture, prioritize data literacy, ensure governance, and target specific business goals (28).

Development of tools like this requires tradeoffs and design choices. We tackled cohort-selection for CECs, but our cohort also includes hundreds of thousands of linked hospitalization records that support research on other chronic disease and clinical phenotypes (6). We leveraged our study-specific and cloud-based data warehouse, but our use of a columnar OLAP database provided a simple option for making large-scale cohort data easily available to a web application. For practical reasons, we omitted our CTS genomic, geospatial, and raw dietary (from two food frequency questionnaires) data from the self-service portion of our platform, but those data can be easily and quickly joined with our platform’s outputs when needed. Some study design components, such as control matching in case-control studies, might be too complex to automate or convert into menu-based choices. We will continue to learn from our user community and improve our CTS Researcher Platform.

## Conclusion

Data providers and data requestors continue to struggle with contemporary cohort selection. Greater use of large-scale survey-based and real world data for research to improve health outcomes will continue to strain today’s manual and labor-intense cohort-selection workflows. The CTS appears to be the first long-running observational cohort to replace its legacy manual cohort-selection methods with a comprehensive web-based self-service application that lets all researchers independently and directly choose, review, receive, and modify the custom data they need for their research. Automated self-service improves efficiency, scalability, and sustainability of data sharing, but ongoing evaluation and community feedback will be essential to identify the right balance of common standards and cohort-specific features and configurations that enable efficient reusability. The CTS Researcher Platform demonstrates that automated and user-friendly self-service cohort selection is practical, even for large and complex data sources, and can be deployed using widely available tools and approaches.

## Data Availability

Based on the original informed consent provided by the California Teachers Study (CTS) participants, CTS data cannot be shared publicly but are accessible to any researcher who registers with the CTS team and agrees to protect the privacy of CTS participants and the confidentiality of all CTS data. Complete details for how to request access to CTS data and resources are available via the CTS website, www.calteachersstudy.org

www.calteachersstudy.org

## Acknowledgements

The collection of cancer incidence data used in the California Teachers Study was supported by the California Department of Public Health pursuant to California Health and Safety Code Section 103885; Centers for Disease Control and Prevention’s National Program of Cancer Registries, under cooperative agreement 5NU58DP006344; the National Cancer Institute’s Surveillance, Epidemiology and End Results Program under contract HHSN261201800032I awarded to the University of California, San Francisco, contract HHSN261201800015I awarded to the University of Southern California, and contract HHSN261201800009I awarded to the Public Health Institute. The opinions, findings, and conclusions expressed herein are those of the author(s) and do not necessarily reflect the official views of the State of California, Department of Public Health, the National Cancer Institute, the National Institutes of Health, the Centers for Disease Control and Prevention or their Contractors and Subcontractors, or the Regents of the University of California, or any of its programs.

The authors would like to thank all California Teachers Study participants and the Steering Committee that is responsible for the formation and maintenance of the Study within which this research was conducted. A full list of California Teachers Study team members is available at https://www.calteachersstudy.org/team.

## References

1. Abrahão MTF, Nobre MRC, Gutierrez MA. A method for cohort selection of cardiovascular disease records from an electronic health record system. Int J Med Inform. 2017 Jun;102:138–149. doi: 10.1016/j.ijmedinf.2017.03.015. Epub 2017 Mar 30. PMID: 28495342.

2. All-of Us Research Program Investigators The All of Us research program. New England Journal of Medicine 381, 668–676 (2019).

3. All of Us / Research Hub / Researcher Workbench. https://www.researchallofus.org/data-tools/workbench/

4. Begg C, Cho M, Eastwood S, Horton R, Moher D, Olkin I, Pitkin R, Rennie D, Schulz KF, Simel D, Stroup DF. Improving the quality of reporting of randomized controlled trials. The CONSORT statement. JAMA. 1996 Aug 28;276(8):637–9. doi: 10.1001/jama.276.8.637. PMID: 8773637.

5. California Teachers Study. www.calteachersstudy.org.

6. California Teachers Study: California Teachers Study Data. https://www.calteachersstudy.org/cts-data

7. California Teachers Study: Researcher Platform. https://calteachersstudy.my.site.com/researchers/s/

8. California Teachers Study: Study Findings. https://www.calteachersstudy.org/study-findings

9. California Teachers Study: Study Population. https://www.calteachersstudy.org/study-population

10. California Teachers Study: Past Questionnaires. https://www.calteachersstudy.org/past-questionnaires

11. Capurro D, Barbe M, Daza C, Santa Maria J, Trincado J. Temporal Design Patterns for Digital Phenotype Cohort Selection in Critical Care: Systematic Literature Assessment and Qualitative Synthesis. JMIR Med Inform. 2020 Nov 24;8(11):e6924. doi: 10.2196/medinform.6924. PMID: 33231554; PMCID: PMC7723741.

12. Conroy MC, Lacey B, Bešević J, Omiyale W, Feng Q, Effingham M, Sellers J, Sheard S, Pancholi M, Gregory G, Busby J, Collins R, Allen NE. UK Biobank: a globally important resource for cancer research. Br J Cancer. 2023 Feb;128(4):519–527. doi: 10.1038/s41416-022-02053-5. Epub 2022 Nov 19. PMID: 36402876; PMCID: PMC9938115.

13. Danciu I, Cowan JD, Basford M, Wang X, Saip A, Osgood S, Shirey-Rice J, Kirby J, Harris PA. Secondary use of clinical data: the Vanderbilt approach. J Biomed Inform. 2014 Dec;52:28–35. doi: 10.1016/j.jbi.2014.02.003. Epub 2014 Feb 14. PMID: 24534443; PMCID: PMC4133331.

14. Data Access Quick Guide to UK Biobank: April 2020. https://md.catapult.org.uk/wp-content/uploads/2020/05/Data-Access-Quick-Guide-UK-Biobank-0420.pdf

15. He T, Belouali A, Patricoski J, Lehmann H, Ball R, Anagnostou V, Kreimeyer K, Botsis T. Trends and opportunities in computable clinical phenotyping: A scoping review. 2023. J Biomed Informatics; 140.

16. He W, Kirchoff KG, Sampson RR, McGhee KK, Cates AM, Obeid JS, Lenert LA. Research Integrated Network of Systems (RINS): a virtual data warehouse for the acceleration of translational research. J Am Med Inform Assoc. 2021 Jul 14;28(7):1440–1450. doi: 10.1093/jamia/ocab023. PMID: 33729486; PMCID: PMC8279787.

17. Horvath MM, Rusincovitch SA, Brinson S, Shang HC, Evans S, Ferranti JM. Modular design, application architecture, and usage of a self-service model for enterprise data delivery: the Duke Enterprise Data Unified Content Explorer (DEDUCE). J Biomed Inform. 2014 Dec;52:231–42. doi: 10.1016/j.jbi.2014.07.006. Epub 2014 Jul 19. PMID: 25051403; PMCID: PMC4335712.

18. Hurst JH, Liu Y, Maxson PJ, Permar SR, Boulware LE, Goldstein BA. Development of an electronic health records datamart to support clinical and population health research. J Clin Transl Sci. 2020 Jun 23;5(1):e13. doi: 10.1017/cts.2020.499. PMID: 33948239; PMCID: PMC8057430.

19. Lacey JV Jr, Chung NT, Hughes P, Benbow JL, Duffy C, Savage KE, Spielfogel ES, Wang SS, Martinez ME, Chandra S. Insights from Adopting a Data Commons Approach for Large-scale Observational Cohort Studies: The California Teachers Study. Cancer Epidemiol Biomarkers Prev. 2020 Apr;29(4):777–786. doi: 10.1158/1055-9965.EPI-19-0842. Epub 2020 Feb 12. PMID: 32051191; PMCID: PMC9005205.

20. Membership of the NCI Cohort Consortium. https://epi.grants.cancer.gov/cohort-consortium/members/

21. Million Veteran Program. https://www.mvp.va.gov/pwa/

22. Murphy SN, Visweswaran S, Becich MJ, Campion TR, Knosp BM, Melton-Meaux GB, Lenert LA. Research data warehouse best practices: catalyzing national data sharing through informatics innovation. J Am Med Inform Assoc. 2022 Mar 15;29(4):581–584. doi: 10.1093/jamia/ocac024. Erratum in: J Am Med Inform Assoc. 2022 Jul 12;29(8):1445. Erratum in: J Am Med Inform Assoc. 2022 Dec 13;30(1):209. PMID: 35289371; PMCID: PMC8922176.

23. PAR-20-294: Core Infrastructure Support for Cancer Epidemiology Cohorts. https://grants.nih.gov/grants/guide/pa-files/PAR-20-294.html

24. Push Button Data Sharing: Web-Based Self-Service and Automated Data Delivery in the California Teachers Study. https://epi.grants.cancer.gov/cohort-consortium/cohort-events.html. Jan 12, 2021.

25. Research Lifecycle. https://researchsupport.harvard.edu/research-lifecycle

26. Rolland B, Geiger AM. Addressing Challenges in Converting Grant-Funded Infrastructures to Broadly Used Research Resources. Cancer Epidemiol Biomarkers Prev. 2019 Oct;28(10):1559–1562. doi: 10.1158/1055-9965.EPI-19-0043. Epub 2019 Aug 28. PMID: 31462397.

27. SEER*Explorer: An interactive website for SEER cancer statistics [Internet]. Surveillance Research Program, National Cancer Institute; 2023 Apr 19. [cited 2023 May 26]. Available from: https://seer.cancer.gov/statistics-network/explorer/. Data source(s): SEER Incidence Data, November 2022 Submission (1975-2020), SEER 22 registries.

28. Self-Service Analytics: How to Use Healthcare Business Intelligence. https://www.healthcatalyst.com/insights/self-service-analytics-how-use-healthcare-business-intelligence

29. Sherlock Cloud Solution & Services. https://sherlock.sdsc.edu/

30. Site Recode ICD-O-3/WHO 2008 Definition. https://seer.cancer.gov/siterecode/icdo3_dwhoheme/

31. Shortreed SM, Cook AJ, Coley RY, Bobb JF, Nelson JC. Challenges and Opportunities for Using Big Health Care Data to Advance Medical Science and Public Health. Am J Epidemiol. 2019 May 1;188(5):851–861. doi: 10.1093/aje/kwy292. PMID: 30877288.

32. Swerdlow AJ, Harvey CE, Milne RL, Pottinger CA, Vachon CM, Wilkens LR, Gapstur SM, Johansson M, Weiderpass E, Winn DM. The National Cancer Institute Cohort Consortium: An International Pooling Collaboration of 58 Cohorts from 20 Countries. Cancer Epidemiol Biomarkers Prev. 2018 Nov;27(11):1307–1319. doi: 10.1158/1055-9965.EPI-18-0182. Epub 2018 Jul 17. PMID: 30018149.

33. The UK Biobank Research Analysis Platform. https://www.ukbiobank.ac.uk/enable-your-research/research-analysis-platform

34. Vander Weele, TJ. Observational Studies and Study Designs: An Epidemiologic Perspective. Observational Studies. 2015;1(1):223–230.

35. von Elm E, Altman DG, Egger M, Pocock SJ, Gøtzsche PC, Vandenbroucke JP; STROBE Initiative. The Strengthening the Reporting of Observational Studies in Epidemiology (STROBE) Statement: guidelines for reporting observational studies. Int J Surg. 2014 Dec;12(12):1495–9. doi: 10.1016/j.ijsu.2014.07.013. Epub 2014 Jul 18. PMID: 25046131.

36. Walters KM, Jojic A, Pfaff ER, Rape M, Spencer DC, Shaheen NJ, Lamm B, Carey TS. Supporting research, protecting data: one institution’s approach to clinical data warehouse governance. J Am Med Inform Assoc. 2022 Mar 15;29(4):707–712. doi: 10.1093/jamia/ocab259. PMID: 34871428; PMCID: PMC8922173.

37. Women’s Health Initiative (WHI) Query Builder. https://www.whi.org/qb/

38. Writing Effective User Stories. https://tech.gsa.gov/guides/effective_user_stories/

